# Acute-stress induced change in salience network coupling prospectively predicts post-trauma symptom-development

**DOI:** 10.1101/2021.07.03.21259969

**Authors:** Wei Zhang, Reinoud Kaldewaij, Mahur M. Hashemi, Saskia B.J. Koch, Annika Smit, Vanessa van Ast, Christian F. Beckmann, Floris Klumpers, Karin Roelofs

## Abstract

Substantial individual differences exist in how acute stress affects large-scale neurocognitive networks, including Salience (SN), Default Mode (DMN) and Central Executive Networks (CEN). Changes in the connectivity strength of these network upon acute stress may predict vulnerability to long-term stress effects, which can only be tested in prospective longitudinal studies. Using such longitudinal design, we investigated whether the magnitude of acute-stress induced functional connectivity changes (delta-FC) predicts the development of posttraumatic stress disorder (PTSD) symptoms in a relatively resilient group of young police students that are known to be at high risk for trauma-exposure.

Using resting-state fMRI, we measured acute-stress induced delta-FC in 190 police recruits before (baseline) and after trauma exposure during repeated emergency aid services (16-month follow-up). Delta-FC was then linked to the changes in perceived stress levels (PSS) and post-traumatic stress symptoms (PCL and CAPS).

Weakened connectivity between the SN and DMN core regions upon acute stress induction at baseline predicted longitudinal increases in perceived stress level but not of post-traumatic stress symptoms, whereas increased coupling between the overall SN and anterior cerebellum was observed in participants with higher clinician-rated PTSD symptoms, particularly intrusion levels. All effects remained significant when controlling for trauma exposure-levels and cortisol stress-reactivity. Neither hormonal nor subjective measures exerted similar predictive or acquired effects.

The reconfiguration of large-scale neural networks upon acute stress induction is relevant for assessing and detecting risk and resilience factors for PTSD. This study highlights the SN connectivity-changes as a potential marker for trauma-related symptom-development, which is sensitive even in a relatively resilient sample.

## Introduction

Exposure to severely stressful events can lead to a wide range of mental health problems, including post-traumatic stress symptoms ^1^. As most individuals with traumatic experiences do not develop significant symptoms in the long term ^2^, an important question is what psychological and biological processes constitute resilience to trauma ^3^. So far, the vast majority of research studying the neural mechanisms underlying stress-related psychopathology has adopted cross-sectional designs. These studies have identified a number of critical brain regions that may be involved in resilience but could also potentially reflect the symptoms that have already arisen ^4^. A smaller number of studies, with generally limited sample sizes, has assessed which brain response prospectively predict later trauma symptom development by taking advantage of longitudinal designs with fMRI scans acquired before trauma exposure ^5^. Interestingly, converging evidence from these studies suggest hyperactive amygdala as a predisposing factor for PTSD ^4–6^. However, these studies have largely used task-based fMRI that is tailored to investigate targeted neural regions of interest, and thus generally leave open the question regarding the involvement of larger scale neural networks, beyond the regional focus ^5^.

Acute stress has been shown to induce a re-allocation of resources from three large-scale neural networks: the salience (SN), default mode (DMN) and central executive network (CEN) ^7,8^. This neural network reconfiguration has been speculated to prioritize resources to facilitate processing of challenging situations in health^7–9^. However, frequent and chronic exposure to stressors likely leads to unfavorable consequences, such as stress-related disorders that have been associated with altered connectivity of SN and DMN^4,10,11^. To date it remains unclear whether the magnitude of brain network reorganization upon short-term challenges is predictive of one’s vulnerability to or resilience against the negative consequences of long-term trauma exposure. Here, using a well-powered prospective longitudinal design, we tested this question in 190 Dutch police recruits who experienced a variety of potentially traumatic events in the line of duty.

Abnormal hyper-connectivity between the core regions of the SN (e.g., amygdala-insula and amygdala-dACC), and hypo-connectivity between the core regions of the DMN (e.g., vmPFC-PCC), as well as between core regions of these two networks (e.g., amygdala-vmPFC) were observed in PTSD patients in contrast to healthy controls^12–15^. In additional to SN and DMN, previous studies also reported decreased CEN connectivity upon acute stress induction in a task condition, and the functional disruptions of the CEN in relation to PTSD ^10,16–21^. It therefore has been proposed that hypoactive DMN and CEN that are overwhelmed by a hyperactive and strongly connected SN may better characterize PTSD ^15^. Yet, due to the nature of cross-sectional designs with post-trauma assessments, these studies cannot inform whether the observed abnormalities existed prior to trauma exposure or were acquired along symptom development. The scarce pioneering longitudinal studies with small sample sizes (e.g., N between 15 to 50)^22–24^ were likely statistically underpowered according to recent guidelines^25,26^ and therefore require replications with large sample sizes. Besides, although these studies provided hints for a linkage between the altered functions of core regions of the SN, DMN, CEN and stress-related disorders, only very few investigations directly deployed network-level approaches.

Since aberrant functional organizations of DMN, SN and CEN have been proposed to underlie a wide range of psychopathologies, including stress-related ones such as depression anxiety and PTSD^10^, it is critical to empirically test whether network-level connectivity profiles in fact can prospectively impact subsequent symptom development after real-life trauma exposure and to elucidate whether these neural correlates can serve as potential risk or resilience factors. Our recent study examining network connectivity changes in response to acute stress induction demonstrated that connectivity changes of the SN (including dorsal anterior cingulate cortex, anterior insula and amygdala) and DMN (including posterior cingulate cortex, precuneus and ventromedial prefrontal cortex) were correlated with cortisol increases after stress induction, respectively^8^. Given the implication of cortisol in inhibiting sympathetic stress responses and regaining physiological homeostasis following acute stressors^27,28^, results from this previous work suggested an adaptive reorganization of brain functional networks in the face of challenges. With these findings, as well as the evidence showing the involvement of the SN, DMN and CEN in stress-related symptoms^11,15^, we asked whether these short-term adaptive responses at the neural network level could predict long-term consequences as a function of trauma exposure. Although stress-induced dynamical changes may be key to the identification of stress-resilience and vulnerability factors ^3^, such changes within and between these large-scale networks have not prospectively been tested in relation to long-term consequences after trauma exposure.

Using a longitudinal design, the current study aimed to elucidate whether the magnitude of acute-stress induced network reconfiguration could predict stress-related symptom development after trauma exposure. Specifically, we used acute stress-induced connectivity changes (i.e., delta-FC) of the SN, DMN, and CEN at baseline (i.e., Wave1 assessment) to *predict* the perceived stress and PTSD symptom levels after continuous exposure to police-operation related trauma in the training period of recruits. We further investigated *acquired* abnormalities in network responses to stress induction after trauma exposure, using neuroimaging data collected at the follow-up assessment (i.e., on average 16 months after the baseline assessment)

Based on the literature, we expected to observe predictive effects of acute stress-induced delta-FC in SN and DMN on trauma-related symptom development. More specifically, the adaptive responses to acute stress that were indicated by increasing SN and decreasing DMN connectivity at the baseline assessment, as shown in our previous study^8^ were expected to be associated with lower levels of stress and PTSD symptoms after trauma exposure. For acquired effects, individuals with higher post-trauma stress levels were expected to show intensified SN connectivity and reduced DMN connectivity after stress induction as suggested in the PTSD literature^12–15^. CEN connectivity changes after acute stress induction were not associated with individual cortisol stress responses in our previous work^8^. However, given prior observations of aberrant CEN connectivity in stress-related psychopathologies ^10,16–21^, we further explored whether acute stress-induced delta-FC of CEN could predict long-term post-trauma stress levels.

## Materials and methods

### Participants

An initial sample of 321 police recruits from Dutch Police Academy participated in the current study in accordance with the principles of the Declaration of Helsinki and with the approval from the Independent Review Board Nijmegen (IRBN), the Netherlands. All participants gave their written informed consent before the study upon their first lab visit (Wave1). Exclusion criteria included any current psychiatric or neurological disorder, history of, or current endocrine or neurological treatment, current use of psychotropic medication, and current drug or alcohol abuse (full details in protocol article^29^). A group of participants (N=86) reported that their core trauma, the trauma most central to their symptoms, had occurred already before our baseline assessment (Wave 1). As we cannot disentangle predisposed and acquired factors for these participants due to the lack of data before Wave 1 assessment, data from these participants were excluded. Further exclusion included data from the participant whose baseline PCL-5 scores met PTSD diagnosis cutoff (i.e., above 33) or who exhibited excessive motion artefacts (i.e., top 5% of participants showing the largest head motion effects from each fMRI session with the mean frame-wise displacement up to 0.39mm)^8^, resulting in a final sample of 190 participants (mean age=23.88) that had complete data from all measurements (see *Figure 1* for detailed sample selection and *Table 1* for demographic information).

**Table 1.** Sample demographics.

| **Sex** | **Sample Size** | **Age (m/sd)** | **Experienced**<br>**Trauma Types**<br>**(m/sd)** |
| --- | --- | --- | --- |
| **Female** | 48 | 23.84/4.67 | 5.04/3.14 |
| **Male** | 142 | 24.0/5.69 | 4.8/3.5 |

**Figure 1.**
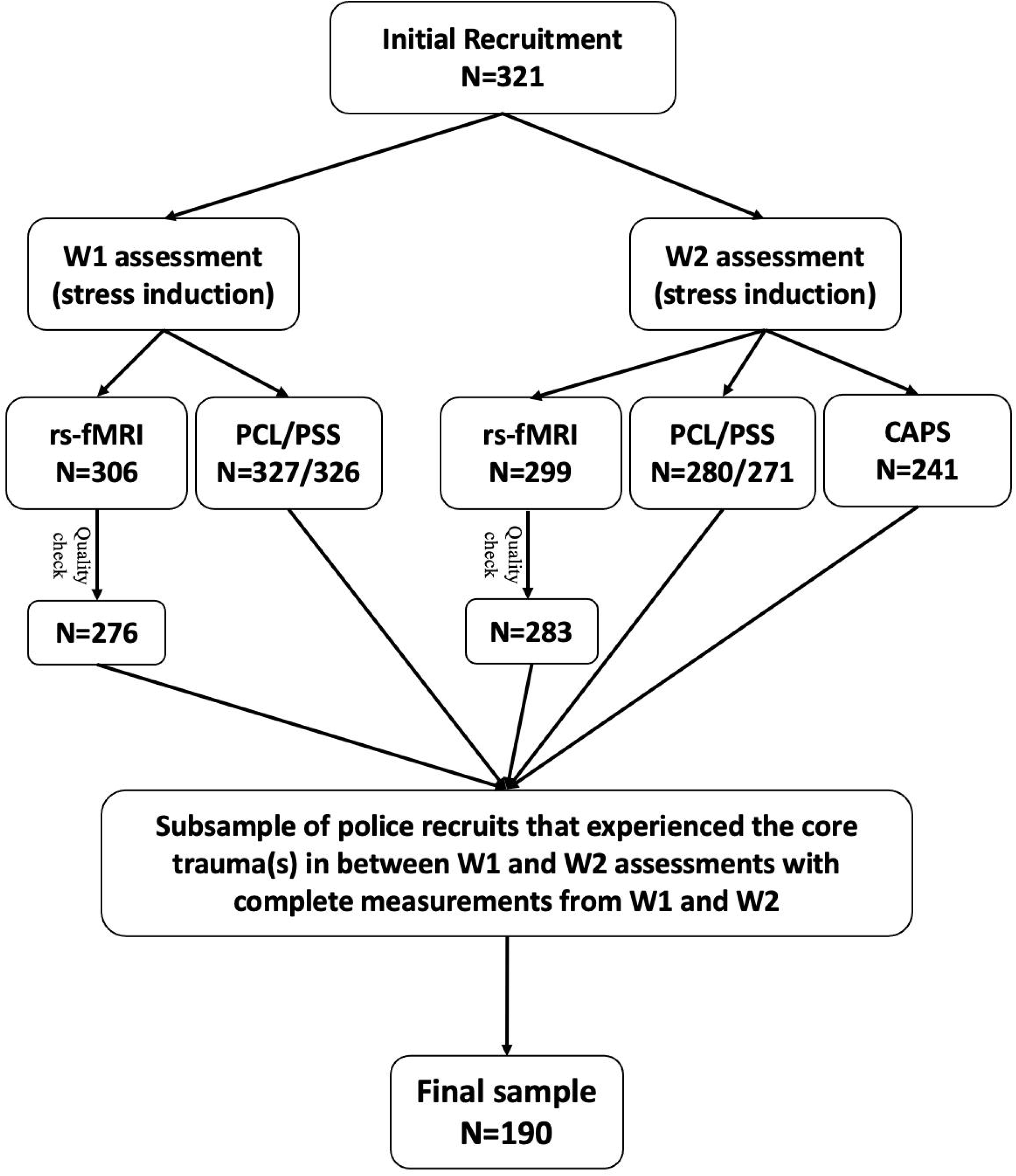
A total number of 321 police recruits participated in the current study. Data from a subsample were acquired for resting-state functional MRI (rs-fMRI) and stress level-related questionnaires (i.e., PTSD Checklist, PCL; Perceived Stress Scale, PSS) or interviews (i.e., Clinician-Administered PTSD Scale, CAPS), at both Wave1 and Wave2 assessments. Following data quality check and the screening of core trauma experiences (i.e., occurrence in between two assessments), data from a final sample of 190 participants were used for further analyses.

### Procedure

The baseline assessment (i.e., Wave1) took place in parallel with the early police curriculum of mostly in-class theoretical trainings. In this assessment, participants filled out questionnaires measuring their baseline levels of perceived stress (using Perceived Stress Scale, PSS) and stress-related symptoms (using PTSD Checklist for DSM-5, PCL-5), prior to the participation in all other experimental tasks. Acute stress induction was conducted in the late afternoon (i.e., between 4-7pm) to ensure stable salivary cortisol levels, which consisted of a SECPT (Socially Evaluated Cold Pressure Task) and MA (mental arithmetic) task (see details in the *Supplemental Materials and Methods*) ^8,30,31^. Hormonal and subjective stress responses were assessed multiple times throughout the experiment (i.e., approximately at -10, 0, +10, +20 and +30 minutes with respect to the experiment onset), and two sessions of resting-state fMRI (rs-fMRI) data with identical acquisition length of 500 scans (367.5s) were acquired immediately before and after stress induction to assess stress-induced functional connectivity changes (*Figure 2*). After an average of 16 months (SD=1.9), participants were tested for the follow-up assessment (i.e., Wave2), when identical measurements were repeated to investigate consequences of exposure to trauma-like events. In the Wave2 assessment, the Police Life Event Scale (PLES)^32^ was additionally used to index the amount of trauma exposure. Participants also participated in a telephone interview comprising the Clinician-Administered PTSD Scale for DSM-5 (CAPS; see full details about all measurements in the protocol article^29^).

**Figure 2.**
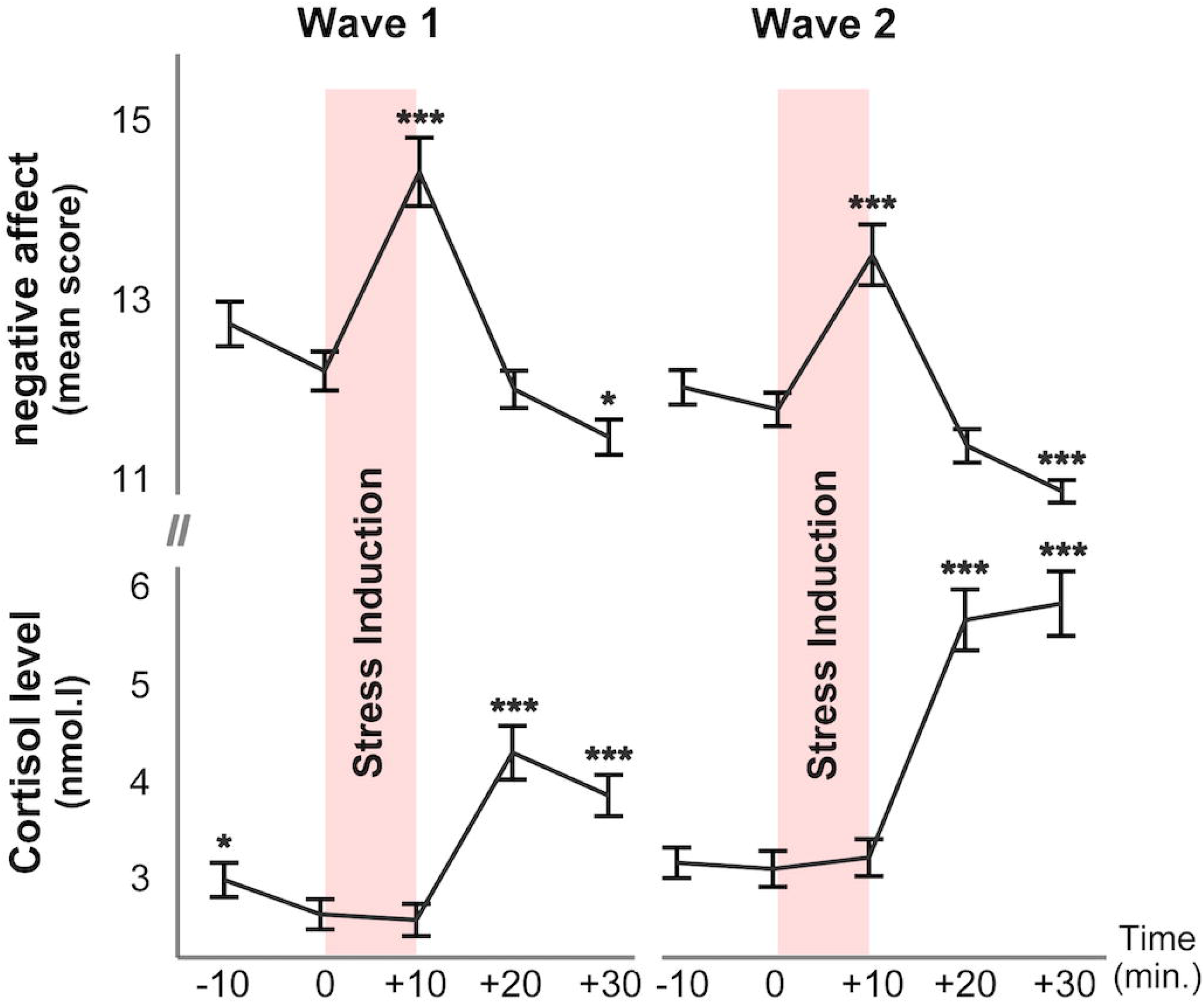
Hormonal and behavioral responses to acute stress induction. Acute stress induction has led to comparable response patterns in hormonal and subjective measures in Wave1 and Wave2 assessments. Negative affect peaked immediately after the onset of stress induction and declined thereafter until eventually below the pre-stress baseline level. Cortisol showed the expected downward trend observed after the morning peak, which was followed by a peak at 20 minutes after the onset of stress induction and remained high. Although overall cortisol increases (i.e., difference between +20 and 0 minutes) were larger in Wave2 than in Wave1 assessments (p<0.05), no differences were observed in negative affect responses (i.e., difference between +10 and 0 minutes). Error bars represent SEM (standard error of measurement) and asterisks indicate significant differences relative to the pre-stress baseline level at time 0 min. ***p<0.0001; *p<0.05.

### Data acquisition and analysis

#### Imaging data acquisition

Acquisition of the resting-state fMRI data occurred before and after the stress induction experiment in both Wave1 and Wave2 assessments, where participants were instructed to lie still and watch a small white cross at the screen center. Imaging data were acquired using a 3T Siemens Magnetom Prisma^fit^ MRI scanner (Erlangen, Germany) with a 32-channel head coil. A multi-band T2*-weighted EPI sequence with acceleration factor 8 (MB8), optimized from the recommended imaging protocols for the Human Connectome Project, was used to acquire a total number of 500 volumes of BOLD-fMRI images (TR=735ms, TE=39ms, flip angle=52°, voxel size=2.4×2.4×2.4mm^3^, slice gap=0mm, FOV=210mm). High-resolution structural images (1×1×1mm^3^) were also acquired, using a T1-weighted MP-RAGE sequence (TR=2300ms, TE=3.03ms, flip angle=8°, FOV=256×256×192mm^3^).

#### Assessment of stress-related measures

Perceived stress level and PTSD symptom levels were measured at both the baseline (Wave1) and follow-up (Wave2), prior to the implementation of the acute stress induction experiment (see Procedure). Additionally, CAPS interviews were conducted at Wave2 assessment. While sum scores of CAPs were used to indicate posttraumatic symptom levels, the change scores of PSS (i.e., Wave1 score subtracted from Wave2 score, delta-PSS) and PCL (delta-PCL) were calculated to indicate the development of posttraumatic stress levels.

#### Assessment of stress-induced hormonal and behavioral measures

To index acute endocrine and subjective stress responses, salivary samples and self-reported ratings of negative affect were measured throughout the stress induction experiment in Wave1 and Wave2 assessments. Following our previous practice^8^, increases in salivary cortisol and negative affect ratings were calculated for each participant to index the magnitude of acute stress responses. Specifically, cortisol increase was defined as the cortisol level 20 minutes after stress induction onset (i.e., at time +20 min. when responses peaked) subtracting baseline level immediately before stress induction (i.e., at time 0 min.). Negative affect increase was calculated as the difference in ratings between the baseline (time 0 min.) and 10 minutes after the onset of stress induction (time +10 min.; *Figure 2*).

#### fMRI preprocessing and analysis

##### Preprocessing

Preprocessing of rs-fMRI data included motion correction, 5mm spatial smoothing, ICA-AROMA based denoising^33^, and high-pass filtering with a cut-off of 100 seconds. Mean signal intensity of white matter and cerebrospinal fluid, as well as head motion parameters were regressed out to minimize psychophysiological confounds and motion artefacts^34,35^. The resulting residual images were subsequently registered to the MNI atlas and used for statistical analyses. Detailed preprocessing can be found in the *Supplemental Materials and Methods*.

#### Identifying delta-FC of resting-state networks

Resting-state networks (RSNs) of interest (i.e., SN, DMN and CEN) were identified from the components of a group-level independent component analysis (ICA) that showed the highest spatial correlation with pre-selected functional ROIs (i.e., anterior SN, left CEN, right CEN and ventral DMN) from the Stanford FIND atlas^8^. Changes in acute stress-induced functional connectivity (i.e., delta-FC) of these RSNs were defined as the connectivity difference between scans pre- and post-stress induction in each of three RSNs (i.e., SN, DMN and CEN). Importantly, we defined the delta-FC at two levels: the local level refers to the delta-FC within each individual RSN, whereas the global level indicates the delta-FC of each RSN with the brain regions also outside the pre-defined networks. Coefficients of both local and global delta-FC were included in the analyses.

#### Testing predictive and acquired effects

To indicate the magnitude of changes in functional connectivity as a function of acute stress induction, coefficients of delta-FC were extracted using the pre-defined local and global networks. Both local and global delta-FC coefficients from Wave1 assessment were used to *predict* stress-related symptom development (i.e., changes in symptoms between two assessments). In contrast, to assess potential acquired changes in network connectivity strength consequential to trauma, we calculated changes of these delta-FC coefficients over time (from Wave1 to Wave2) and looked at the associations with symptom development.

To obtain the local level delta-FC coefficients, we used the RSNs maps derived from group-level ICA with a threshold of Z>3 to reduce noise while retaining the overall spatial patterns of each network. The changes in the beta values within the resulting mask were then used to index the network connectivity changes after stress induction. For the more global level delta-FC coefficients, we calculated the differential RSNs at the individual level (i.e., derived from dual-regression analysis) before and after stress induction (i.e., subtraction between two scans), thresholded at p<0.167 and averaged the resulting maps across participants. This essentially created a group-level map of regions showing consistent changes in connectivity with the network of interest after stress induction. Changes in the averaged beta values across all these regions were taken as a summary measure of stress-related global connectivity changes for that network (also see *Supplemental Materials and Methods*) ^8^. This two-level network approach allowed us to examine for each individual RSN upon the perturbation by acute stress induction the internal communications among nodes (i.e., local level delta-FC), as well as for their interplays with all brain areas that showed a connectivity change with the network (i.e., global level delta-FC).

Additionally, as cortisol stress reactivity was previously found predictive of subsequent PTSD symptom development in a limited number of studies^36,37^, we tested whether this could be replicated using acute stress-induced cortisol increases at baseline to predict stress-related symptomatology in our study. We further examined the changes in cortisol increase and negative affect upon acute stress induction as a function of trauma exposure to explore whether symptom changes are companied by hormonal and behavioral changes (i.e., acquired effects).

Delta-PSS, delta-PCL and CAPS sum scores were calculated to indicate each individual’s posttraumatic stress levels. These change scores were used as the outcome measures in our analyses for predictive and acquired effects of symptom development. We further explored the development of specific symptom clusters, using the sum scores of each sub-cluster in delta-PCL and CAPS measures (*Table S1*).

### Statistical analyses

Spearman rank correlation was used for all correlation analyses to mitigate the influences from extreme values and reduce the chance of false positives. Concerning the results for our a-priori hypotheses (i.e., regarding the delta-FC of SN and DMN), FDR corrections were applied to account for the number of analyses involving three outcome measurements (i.e., delta-PCL, delta-PSS and CAPS scores). For more exploratory analyses concerning the delta-FC of CEN for which we had no a priori hypotheses, more stringent FDR corrections were considered to account for two levels of network connectivity (i.e., local and more global levels) and three outcome measurements. Follow-up tests on sub-cluster symptom scores were carried out only if predictive or acquired effects were observed. Concerning these analyses, FDR corrections were conducted to account for the number of analyses involving all four sub-cluster symptom scores, for the local and more global level connectivity, separately. In case of significant results concerning delta-PCL or delta-PSS (either the overall changing score or the sub-cluster score), semi-partial Spearman correlation was further conducted to control for the baseline PCL or PSS level from Wave1 assessment. Finally, we used a Generalized Additive Model (GAM) to explore whether the predictive effects of hypothesized neural measures remain significant when accounting for influences of stress reactivity at hormonal and behavioral levels from baseline assessment, and the impact of trauma exposure (i.e., indicated by the number of experienced trauma types). Similar analyses were conducted for acquired effects to account for changes in effects of cortisol and negative affect as a function to acute stress induction between two assessments. Unlike multiple linear regression that estimates a single parameter for each predictor, GAM finds unspecified (non-parametric) functions that relate the predicted Y (dependent variable) values to the predictor values, and thus allows non-parametric fit^38,39^.

Apart from the coefficient extraction that was carried out using FSL^40^, all other statistical analyses were conducted using R version 3.6.1^41^, with *pcor* function from RVAideMemoire package^42^ and *gam* function from mgcv package^43^ specifically for running semi-partial correlation and GAM analyses, respectively.

## Results

### Acute stress responses

Successful acute stress induction was observed in both baseline (Wave1) and follow-up (Wave2) assessments. Specifically, increases in salivary cortisol and reported negative affect were observed in Wave1 following stress induction, as reflected in main effects of sampling time (F_cortisol_(4, 677.36)=76.82, p<0.0001; F_affect_(4,719.87)=51.50, p<0.005). Similar significant effects were observed in Wave2 (F_cortisol_(4,644.18)=123.4, F_affect_ (4,675.06)=51.03, p’s<0.0001). In short, our experimental manipulation successfully induced acute stress, indicated by increases in cortisol and negative affect for both waves (see *Figure 2*).

### Traumatic experiences and posttraumatic stress measures

In between our two waves of data collection, the police recruits on average experienced 6.63 different types of potentially traumatic events (SD=3.78) with a range between 0 and 17. Most frequently experienced trauma were encountering suicide (including attempt; 31.4%), severe (traffic) accidents (23.6%) and physical assault (17.8%; *Figure S1*).

From Wave1 to Wave2, average stress symptom levels slightly increased with no statistically significant changes at the group level in either perceived stress (PSS: t(182)=1.48, p=0.14) or overall PTSD symptom levels (PCL: t(189)=0.77, p=0.44). Nevertheless, closer inspection revealed large variance in individual symptom trajectories (*Figure 3; Table S2*), together with a significant increase in intrusion symptom level (t(189)=2.22, p<0.05; other symptom clusters p>0.05). At Wave2, the average clinical interview (CAPS) score of overall PTSD symptoms was 1.79 (SD=4.01; sum score range: 0-27), with three participants having developed full-blown PTSD according to DSM-5 criteria (*Figure S2*). Although the group-level CAPS score was relatively low, a substantial proportion of police students (about 80%) exhibited clinically relevant increases in symptom levels (i.e., reported at least one symptom in each cluster in Wave2 PCL). We therefore proceeded to test whether the large variation in PTSD symptom trajectories could be explained by the stress-related neural network connectivity changes.

**Figure 3.**
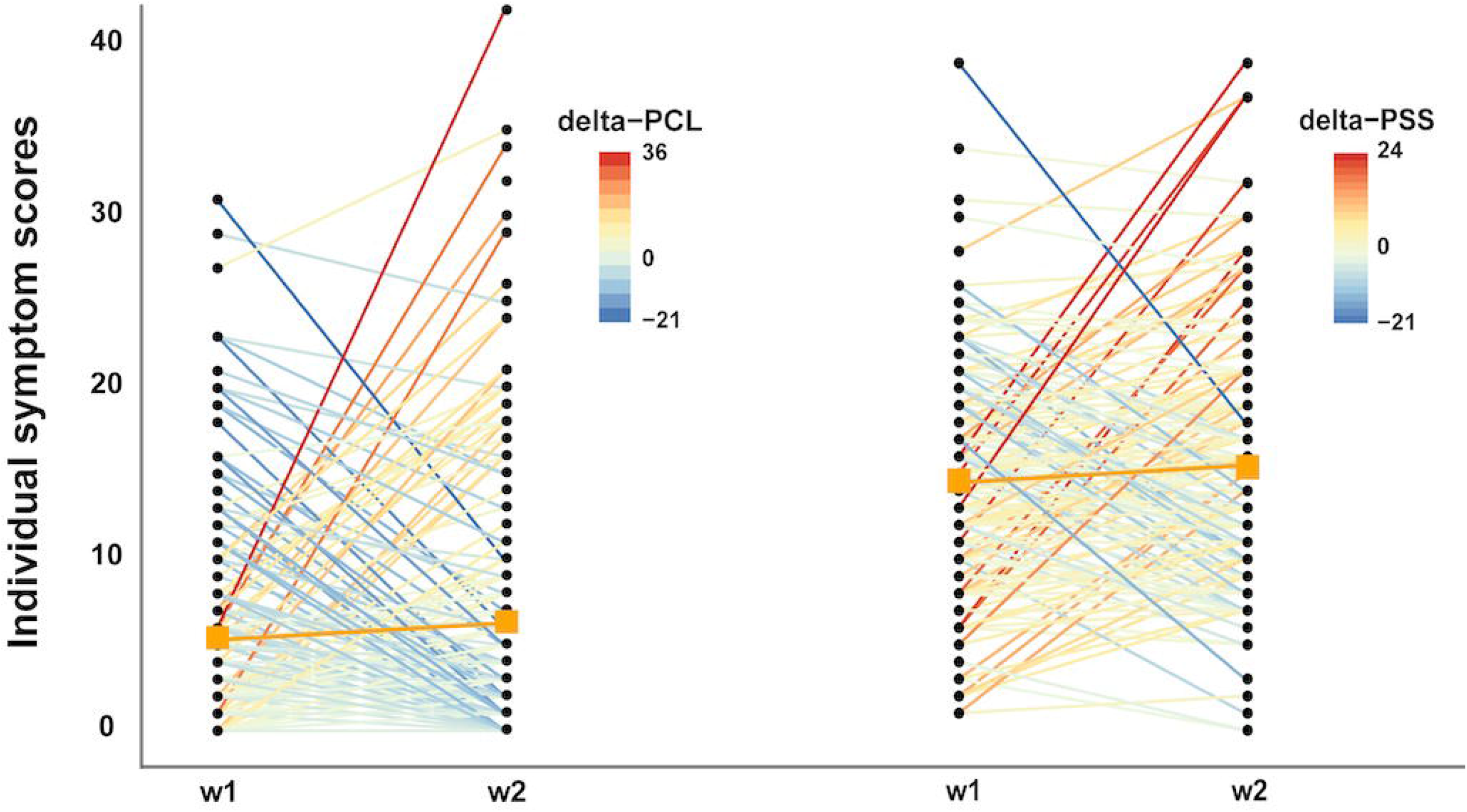
Development of PTSD symptom levels (assessed by PCL-5) and perceived stress levels (assessed by PSS) from Wave1 (w1) to Wave2 (w2). Large individual differences were observed for change scores in PCL (i.e., delta-PCL, left panel; range: -21 to +36), and in PSS (i.e., delta-PSS, right panel; range -21 to +24). Each line represents the change of individuals in stress symptoms from w1 to w2. Group means for both PCL and PSS from each wave assessment are illustrated using the orange squares.

### Predictive effects of baseline acute stress responses for subsequent trauma symptom development

In testing our a prior hypotheses concerning the SN and DMN, a decreased coupling between SN and DMN core regions (i.e., posterior cingulate cortex/precuneus) following stress induction at baseline was found predictive of larger increases in perceived stress level after trauma exposure (rho=-0.19, p=0.0094; *Figure 4*). This effect remained significant after FDR correction for multiple comparisons (p_fdr_=0.0167) and when baseline level PSS was controlled for (rho=-0.19, p=0.0039). This predictive effect of SN-DMN coupling remained as the only significant predictor in our follow-up analysis using Generalized Additive Model (GAM) when influences of baseline cortisol and subjective affect, as well as trauma exposure amount were considered (F=7.20, p=0.008). Against our hypotheses, we observed no predictive effects for self-reported PTSD symptoms (delta-PCL) or clinician rated PTSD levels (CAPS scores), nor with respect to hypothesized DMN connectivity changes after stress induction (all uncorrected p’s>0.08).

**Figure 4.**
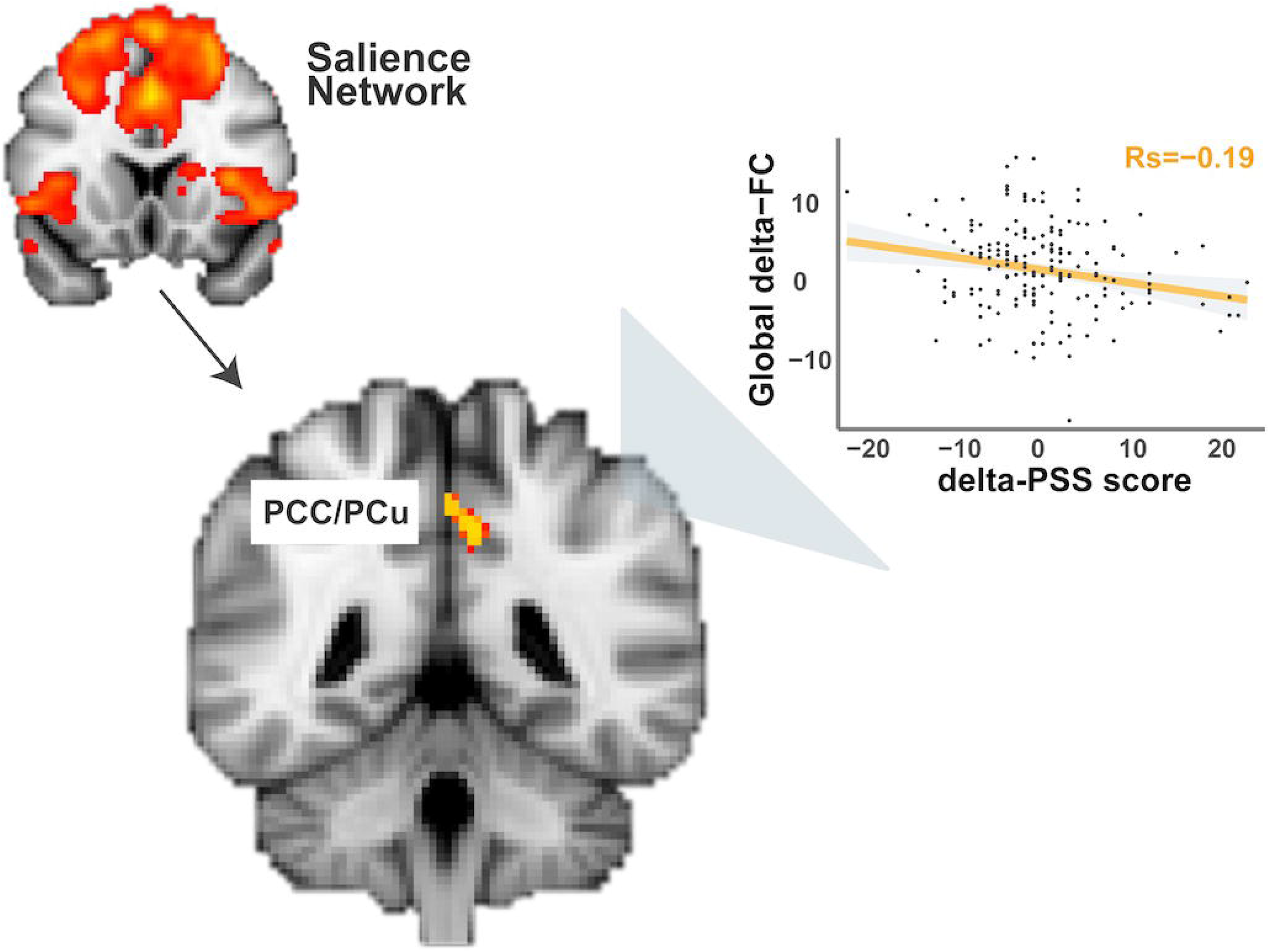
Decreases in the coupling between the overall SN and hub regions of the DMN (i.e., posterior cingulate cortex (PCC)/precuneus (PCu), and postcentral gyrus and intracalcarine cortex as a function of acute stress induction predicted the higher levels of perceived stress after trauma exposure. The arrow indicates functional connectivity changes between the overall salience network (i.e., coefficients extracted using the depicted network mask thresholded at Z>3) and clusters in PCC/PCu, visual cortex and somatosensory cortex.

Subsequent exploratory analyses for CEN revealed that higher clinician-rated PTSD symptoms (i.e., CAPS total score) were predicted by increased delta-FC, both at the local (i.e., delta-FC within the CEN; rho=0.21, p=0.0031) and more global levels (i.e., delta-FC of CEN with brain regions also outside the network; Rs=0.19, p=0.0089). After FDR corrections, only the effect of local CEN delta-FC remained significant (p_fdr_=0.019). Follow-up tests on the sub-cluster symptoms revealed that delta-FC within CEN predicted levels of alteration in mood and cognition (rho=0.19, p=0.0085), as well as hyper-arousal symptoms (rho=0.25, p=0.00058). These effects also remained significant after multiple comparison correction (p_fdr_<0.035; *Figure S3*).

In comparison to these baseline neural responses to acute stress induction, we did not find cortisol reactivity, nor negative affect, predictive of stress-related symptomology development (all p’s>0.05).

### Acquired effects after trauma exposure

C*hanges* in acute stress responses from Wave1 to Wave2 at hormonal, behavioral and neural levels were linked to increases in symptomology to test for acquired abnormalities. Although an increase was observed at the group level in cortisol stress responses from Wave1 to Wave2 assessments (t(150) = 3.52, p<0.001), we did not find any associations between this increased cortisol stress response and symptom level changes (all p’s>0.05). No difference in negative affect levels upon acute stress induction was found between Wave1 and Wave2 assessments, nor did we observe any significant associations between negative affect and symptom change (all p’s>0.05). Furthermore, we did not observe significant associations between connectivity changes of DMN or CEN and symptom changes (all p’s>0.05). Furthermore, there were no significant associations between connectivity changes of DMN or CEN and symptom changes (all p’s>0.05). However, we did observe an increased coupling between the overall SN and anterior cerebellum (i.e., increased delta-FC as a function of acute stress induction) from Wave1 to Wave2 that was associated with higher PTSD symptom levels (i.e., CAPS total score; rho=0.-18, p=0.019). Yet, this effect just missed significance when correcting for multiple comparisons (p_FDR_=0.057). Follow-up tests examining sub-cluster symptoms suggested that this effect might have been driven by intrusion symptom (rho=-0.22, p=0.0038; other symptoms p>0.05), with the participants showing higher intrusion symptom levels also exhibiting larger neural coupling in response to acute stress induction (*Figure 5*). The effect for sub-cluster symptom also remained significant after FDR correction (p_FDR_=0.015), and after accounting for the influence of changes in cortisol and negative affect between two assessments (F=6.05, p=0.015). No significant effects were observed for delta-PCL, nor delta-PSS scores.

**Figure5.**
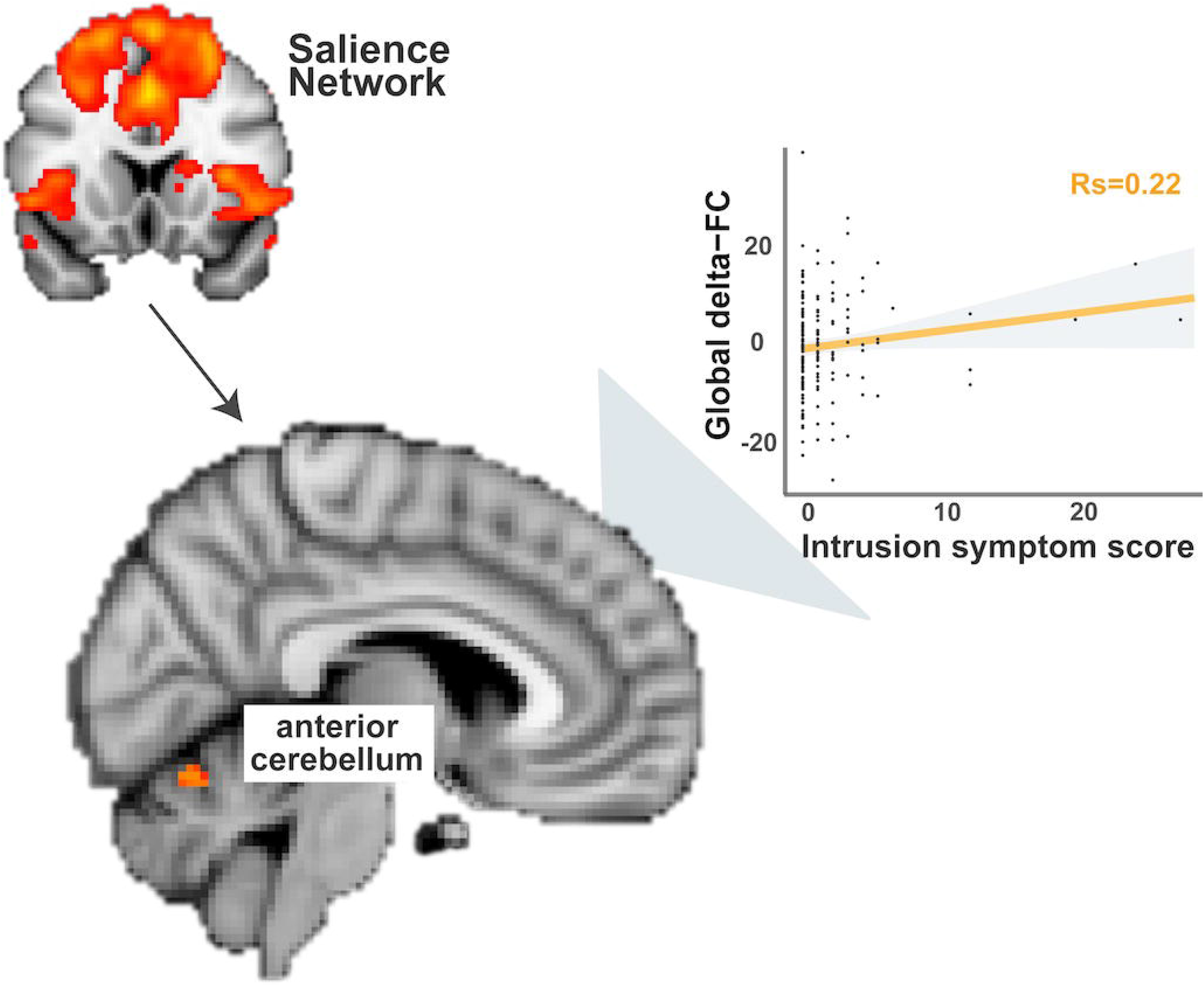
Increased coupling (stemming from reduced decoupling) between the overall SN and anterior cerebellum (as depicted in the brain image) from Wave1 to Wave2 in response to acute stress induction was associated with higher levels of intrusion symptom, indicated by CAPS scores. The arrow demonstrates functional connectivity changes between the overall salience network (i.e., coefficients extracted using the depicted network mask thresholded at Z>3) and clusters in anterior cerebellum. To note, spearman rank correlation was conducted to minimize the influences from the extreme values.

## Discussion

In this prospective longitudinal study, we investigated whether acute stress-induced neural network changes could function as a risk factor to or a resilience factor against the development of PTSD symptoms. To this end, we tested the predictive effects of such changes on long-term stress-related symptomatology after exposure to real-life trauma in police recruits. Reduced global connectivity of the SN with DMN in response to baseline acute stressors predicted increased post-trauma stress levels 16 months later. A different pattern emerged for neural network changes between assessments that followed symptom development and thus appear *acquired* rather than a pre-trauma risk factor: individuals with higher levels of PTSD intrusions symptoms at follow-up showed increased coupling between the SN and anterior cerebellum after acute stress induction in the follow-up versus baseline assessment. Interestingly, both acquired and predictive neural effects in our study were found significant above and beyond the contribution of hormonal and subjective stress measures. Together, these findings suggest the SN connectivity patterns as a function of acute stress may serve as a potential risk factor to the development of stress-related symptoms upon trauma exposure.

To our knowledge, this is the first longitudinal study that has used a network-based approach to link stress-induced connectivity changing patterns of large-scale brain networks at baseline to the subsequent symptom development after trauma exposure. Our findings therefore provide new empirical evidence that the magnitude of large-scale network reconfiguration upon acute stress exposure is relevant for investigating resilience and risk factors for stress-related symptomatology.

In line with our predictions, acute stress-induced decreases in overall SN connectivity with brain regions (i.e., PCC and precuneus) predicted higher perceived stress level after trauma exposure. Acute stress has been shown to immediately prompt SN engagement at the potential cost of neural resources that would otherwise have been allocated to other brain circuits^7,8^. This stress-induced reconfiguration of brain function is hypothesized to facilitate the coping with the challenging situations at hand by reallocating neural resource towards the SN for attention direction towards evolutionary relevant stimuli and integration of top-down appraisal and bottom-up visceral and sensory information (see review by Uddin^44^). Insufficient SN involvement in response to acute stress therefore may signal suboptimal processing such that its dynamic coordination with other brain networks becomes diminished and thus results in undesirable long-term consequences, such as the observed increases in stress levels after exposure to real life trauma. Notably, hyperactivity of the amygdala and hypoactivity of the prefrontal cortex have consistently been implicated in stress-related psychopathology (see review by Fenster et al. ^1^), including our previous work where diminished aPFC control over the amygdala during an approach and avoidance task predicted subsequent PTSD symptoms ^6^. Yet, in the current study, more local level SN connectivity with extracted coefficients also containing signals of bilateral amygdala did not show any predictive effects. This discrepancy is likely due to different study designs: involvement of fronto-amygdala circuit is often seen in task conditions requiring repeated regulation of a series of task-related stimuli, suggesting complimentary insight into PTSD biomarkers provided by different study designs.

Interestingly, SN reconfiguration upon acute stress induction seemed to not only signal a risk factor for later symptom development, but also indicate acquired abnormalities that were associated with increasing symptom levels. Our finding of SN-cerebellum coupling in participants with relatively high PTSD symptoms is in line with a growing number of studies that has linked cerebellum to emotional processing and regulation, particularly to negative emotional memories^45–47^, as well as to pathophysiology of PTSD^48–52^.

Furthermore, our exploratory analyses for CEN found that acute stress-induced increases in connectivity of this network prior to trauma exposure could predict the elevated levels of overall post-trauma symptoms, and of specific negative mood/cognition and hyperarousal symptoms (Figure S3). These results suggest that the reconfiguration of CEN upon acute stressors may allow the tracking of symptom development after trauma exposure. Our findings are in line with the role of dorsal lateral prefrontal cortex (dlPFC), a core region of the CEN in emotion regulation. They are also consistent with the evidence from simulation studies that the modulation of dlPFC activity and connectivity could be beneficial in alleviating PTSD symptoms^53,54^.

Contrary to our prediction, however, we did not observe any associations between acute stress induced delta-FC of DMN at baseline and long-term consequences of trauma exposure. However, the finding of decreased connectivity between SN and posterior DMN suggests that the regions in DMN may function distinctively in response to stressful events over time, hence the mean coefficient indicating overall connectivity patterns of all regions within the network at baseline could not capture individual variability in longitudinal symptom development. Additionally, we did not find evidence in support of cortisol reactivity predicting PTSD symptom development, which was reported in a few recent longitudinal studies^37,55^. Discrepancy may arise from differences in sample characteristics (i.e., combat solders vs. police recruits), analytical approaches (i.e., subtyping vs. continuous modelling), and the timing of assessment with regard to trauma exposure (i.e., once a year for four years vs. twice with 16 months in between) between the previous and our studies. Future investigations that study cortisol stress response in relation to symptom development should consider these differences.

In an attempt to investigate the predictive factors for PTSD symptom development and to disentangle predictive from acquired effects of the neural networks, we focused on a relatively healthy and resilient sample, whose baseline levels of depression, anxiety, and PTSD symptom scores at Wave1 assessment were significantly lower in comparison to a group of age and sex matched control participants (all p’s<0.05). Notably, these observations were in line with the fact that all police students were prescreened for the enrollment based on their physical and psychological performance, and with the literature suggesting higher resilience in police than in civilians ^56^. With limited variations in stress-related psychopathology in the current study sample, our study differs from most of the existing longitudinal (and cross-sectional) studies that have focused on clinical populations with maximized variances pertinent to the symptomology^12,57^. However, our sample here is not affected by the typical confounds that surround more severe psychopathology either (e.g., medication intake). Our dimensional approach therefore potentially allows more direct interpretations of the prediction findings and our resilient sample sheds important light on how stress resilience versus vulnerability might be instantiated in the brain. Further, an important advantage of this study was the leverage of a well-established acute stress challenge in combination with a relatively large sample size.

In conclusion, the current study used connectivity changes of large-scale neural networks in response to an acute stress challenge to predict subsequent stress-related symptoms after trauma exposure in police recruits. Whereas SN-DMN connectivity prospectively predicted the longitudinal changes in perceived stress level, increased SN-cerebellum connectivity was acquired in participants with higher PTSD symptom levels. These findings suggest that acute stress-induced SN connectivity changes may serve as a potential marker of PTSD vulnerability.

## Data Availability

Derived data supporting the findings of this study are available from the corresponding author [WZ] on request.

## Acknowledgements

This work was supported by a VICI grant (#453-12-001) from the Netherlands Organization for Scientific Research (NWO) and a consolidator grant from the European Research Council (ERC_CoG-2017_772337) awarded to Karin Roelofs. We are particularly grateful to Dutch Police Academy (Politieacademie) for their cooperation. We also thank our former and current colleagues Ingrid Kersten, Naomi de Valk, Geoffrey Bertou, Leonore Bovy, Iris Hulzink, Tiele Dopp, Marijolein Hartgerink, Bart Becker, Madine Zoet, Delphin van Benthem, Job de Brouwer, Lisanne Nuijen, Pepijn van Houten, Klaas van Groesen and Nienke Flipsen for their help in setting up the study, recruiting participants and acquiring data, and to Paul Gaalman for his technical assistance in fMRI data acquisition.

