## Supplemental Materials and Methods for "Acute-stress induced change in salience network coupling prospectively predicts post-trauma symptom-development" (from another version of this preprint: version 1, 2021-07-07)

*Procedure*

The experiment of acute stress induction was conducted in the late afternoon, following the questionnaire session in the morning and a variety of other experimental assessments (see full testing schedule in our study protocol^1^). This experiment started with a SECPT (Socially Evaluated Cold Pressure Task), where participants were instructed to first put their right foot in the icy cold water (1-3 ºC) for 3 minutes. Immediately after the SECPT, participants were instructed to count from an odd number (i.e., 2053) backwards as quickly and accurately as possible, in steps of 17 in a MA (mental arithmetic) task. Whenever they made a mistake, they were required to start over with a new number until three-minute time was up^2^. Assessment of stress responses was carried out at hormonal, behavioral and neural levels.

*Preprocessing*

Analysis of fMRI data was performed with FSL5.0.9 (FMRIB, Oxford, UK). The first five images of each resting-state scan were discarded to allow for T2^*^ equilibration effects. Further preprocessing included motion correction, spatial smoothing with a 5mm FWHM kernel, denoising using ICA-AROMA^3^, and high-pass filtering with a cut-off of 100 seconds. To further minimize motion and psychophysiological confounds after the denoising procedure, the six realignment parameters, their temporal derivatives and the quadratic terms of both the original parameters and derivatives were used as motion parameters in a multiple linear regression model^4–6^. Additionally, each individual T1 image was segmented for subject-specific white matter and CSF masks that were subsequently thresholded with a 95% probability and registered with each individual participant’s functional image. Mean signal intensities of white matter and CSF were extracted and included in the regression model^4,7^. The resulting residuals were normalized with standard MNI atlas and analyzed subsequently. For each participant, one pre- and one post-stress rs-fMRI recording were preprocessed and analyzed.

*Network Definition and Extraction of Connectivity Coefficients*

The resting-state networks (RSNs) of interest was defined using Stanford FIND Atlas^8^. Specifically, components from a group-level independent component analysis (gICA) were correlated to FIND Atlas and those showing the highest spatial correlation with the pre-selected functional ROIs (i.e., anterior SN, left CEN, right CEN and ventral DMN) from the FIND Atlas were identified as salience, default mode and central executive networks (SN, DMN, CEN)^2^.

Two levels of delta-FC were utilized in the analyses of the current study: the local and global levels. Importantly, delta-FC here refers to the differences in connectivity between scans before and after stress induction. The local level delta-FC was defined as the mean coefficients extracted from the group-level ICA (gICA) component corresponding to the SN, DMN and CEN, the difference in which between Wave1 and Wave2 assessments was used to indicate the changes of within-network connectivity as a function of acute stress induction over time (i.e., from prior to post trauma exposure). More global-level delta-FC was extracted using the group-level average maps of individual delta-FC images (i.e., the average of connectivity differences in individual level RSNs between scans before and after stress induction). The changes of this whole-brain connectivity (i.e., changes in delta-FC) were used to indicate the coupling between the networks and other brain regions as a function of acute stress induction over time. To ensure the inclusion of only the core regions of each network, the templates used to extract the local-level connectivity coefficients of RSNs were threholded at Z>3 (i.e., on the gICA components). For the templates used to extract the global-level connectivity coefficients, a threshold of p<0.167 was considered to ensure the inclusion of voxels that were statistically significant with respect to the connectivity changes before and after stress induction (i.e., Bonferroni correction adjusted p value).

**Supplemental Results**

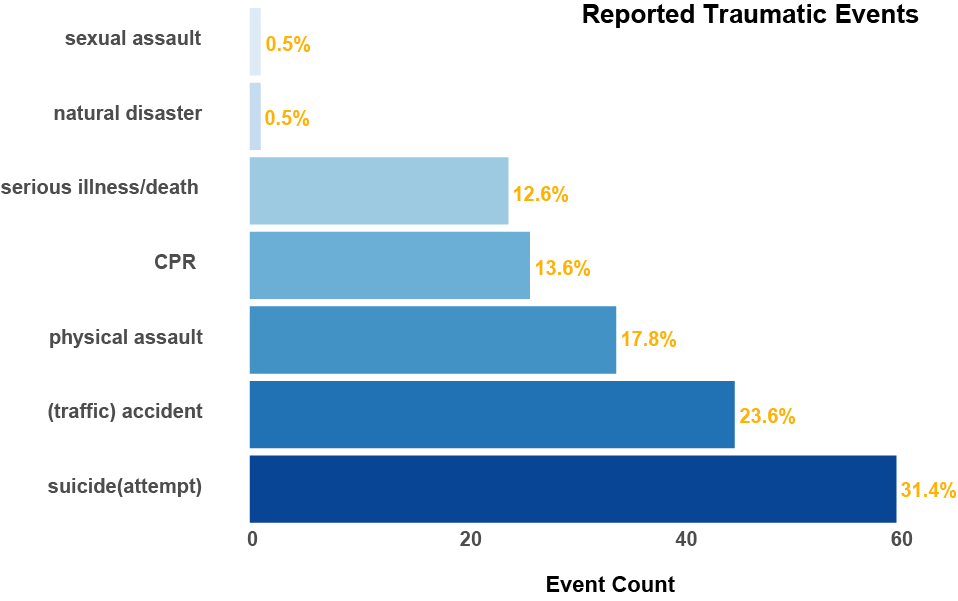

*Figure S1. Counts and frequency (percentage) of reported traumatic events in Clinician Administered PTSD Scale (CAPS) that participants had experienced or witnessed during their emergency aid training (i.e., in between Wave 1 and Wave 2 assessments). Overall, suicide (including attempt) and (traffic) accidents were the most frequently experienced events, followed by physical assault, CPR (cardiopulmonary resuscitation) and serious illness or death. Experiences in natural disasters and sexual assault were most infrequent.*

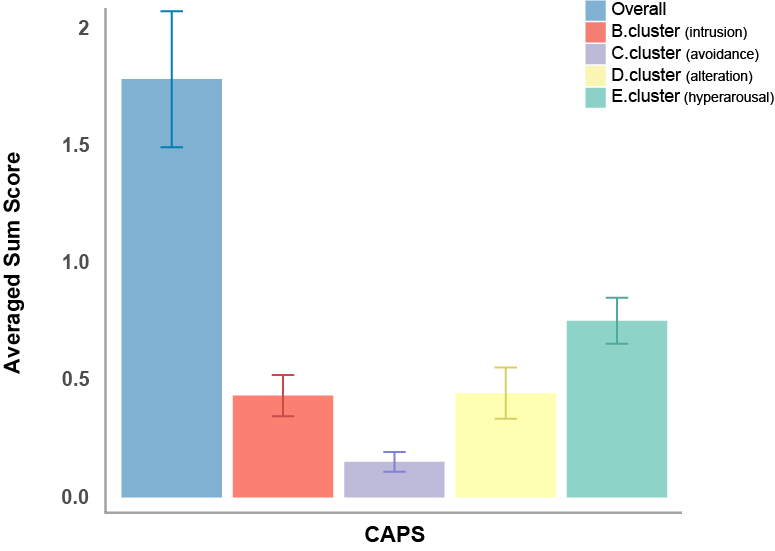

*Figure S2. Averaged sum scores of overall CAPS and each individual sub-cluster symptoms. At group level, overall CAPS scores were significantly higher than zero with increases in each individual sub-cluster symptoms.*

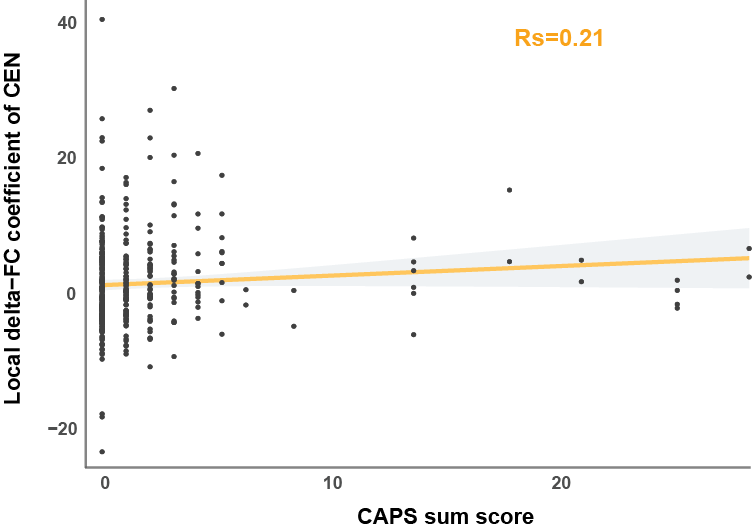

*Figure S3. Increases of connectivity within the CEN predicted higher levels of PTSD symptoms, indicated by CAPS sum scores.*

Table S1. Sample demographics.

| Sex | Size | Age (m/sd) | Experienced Trauma Types (m/sd) |
| --- | --- | --- | --- |
| Female | 48 | 23.84/4.67 | 5.04/3.14 |
| Male | 142 | 24.0/5.69 | 4.8/3.5 |

Table S2. Mean/SD score of outcome measures.

| Measures | | Mean | SD |
| --- | --- | --- | --- |
| delta-PSS | Overall | 0.78 | 7.13 |
| delta-PCL | Overall | 0.62 | 8.85 |
|  | Intrusion (cluster B) | 0.48 | 2.66 |
|  | Avoidance (cluster C) | 0.03 | 1.60 |
|  | Negative alterations in cognition and mood (Cluster D) | 0.09 | 3.60 |
|  | Arousal/reactivity (Cluster E) | 0.02 | 3.10 |
| CAPs | Overall | 1.79 | 4.01 |
|  | Intrusion (cluster B) | 0.43 | 1.22 |
|  | Avoidance (cluster C) | 0.15 | 0.58 |
|  | Negative alterations in cognition and mood (Cluster D) | 0.45 | 1.51 |
|  | Arousal/reactivity (Cluster E) | 0.75 | 1.36 |
